# Prolonged presence of replication-competent SARS-CoV-2 in mildly symptomatic individuals: A Report of 2 Cases

**DOI:** 10.1101/2020.11.18.20232546

**Authors:** Maria Cassia Mendes-Correa, Fabio E. Leal, Lucy S. Villas-Boas, Steven S. Witkin, Anderson de Paula, Tania Regina Tozetto-Mendonza, Noely Evangelista Ferreira, Gislaine Curty, Pedro Santos de Carvalho, Lewis F. Buss, Silvia F. Costa, Flavia Mendes da Cunha Carvalho, Joyce Kawakami, Noemi Nosomi Taniwaki, Heuder Paiao, Joao Carlos da Silva Bizário, Jaqueline Goes de Jesus, Ester Cerdeira Sabino, Camila Malta Romano, Regina Maura Zetone Grepan, Antonio Sesso

**Affiliations:** Faculdade de Medicina da Universidade de Sao Paulo-Laboratorio de Investigacao Medica em Virologia (LIM52)-Instituto de Medicina Tropical de Sao Paulo; Faculdade de Medicina da Universidade Municipal de Sao Caetano do Sul; Instituto Nacional do Cancer-Rio de Janeiro; Weill Cornell Medicine, USA; Departamento de Molestias Infecciosas e Parasitarias da Faculdade de Medicina da Universidade de São Paulo; Faculdade de Medicina da Universidade de Sao Paulo- Instituto de Medicina Tropical de Sao Paulo; Instituto do Coracao do Hospital das Clinicas da Faculdade de Medicina da Universidade de Sao Paulo; Instituto Adolf Lutz de São Paulo

## Abstract

It has been estimated that individuals with COVID-19 can shed replication-competent virus up to a maximum of twenty days after initiation of symptoms. This report describes two patients with mild forms of the disease who shed replication-competent virus for 24 and 37 days, respectively, after symptom onset.

## Introduction

Accumulating evidence indicates that during the COVID-19 pandemic, SARS-CoV-2 RNA can initially be identified in infected individuals 1-3 days before symptom onset (1,2,3). Viral load, as measured by RT-PCR, reaches its highest level during the first week of symptom onset, followed by a gradual decline over time. It has been estimated that replication-competent virus is no longer present in COVID-19 patients 20 days after onset of symptoms (3-8). The majority of studies that addressed this situation involved hospitalized individuals and those with severe disease.

Studies to address the possible presence of SARS-CoV-2 during the different phases of COVID-19 disease in mildly infected individuals, and utilization of viral culture techniques to identify replication-competent viruses, have been limited. This report describes two SARS-CoV-2-infected women with mild disease in which virus, shown to be replication-competent, persisted for longer periods of time than has been reported previously.

## Setting

The described cases were participants in *The Corona São Caetano Program*, a primary care initiative offering COVID-19 care to all residents of São Caetano do Sul, Brazil. Briefly, residents of the municipality with symptoms consistent with COVID-19 were encouraged to contact the Corona São Caetano platform via the website (access at https://coronasaocaetano.org/) or by phone. Participants were invited to complete an initial screening questionnaire that included information on type, onset and duration of symptoms. Those meeting the suspected COVID-19 case definition were then called by a medical student to complete a risk assessment. Individuals meeting pre-defined triage criteria for mild disease were offered a home visit in which a self-collected nasopharyngeal swab was obtained for analysis.

## Ethics

The study was approved by the local ethics committee (Comissão de Ética para Análise de Projeto de Pesquisa - CAPPesq, protocol No. 13915, dated June 03, 2020). The committee waived the need for informed consent and allowed the development of an analytical dataset with no personal identification for the current analysis.

## Clinical Cases

Case 1 is a woman, in her 50’s, whose first contact was in the middle of April, 2020. She reported that 20 days previously, she first experienced a dry cough, headache, asthenia, arthralgia and myalgia. She denied having a fever. Twenty-two days after the onset of symptoms, a nasopharyngeal swab tested positive for SARS CoV-2 RNA. Subsequently, she developed nausea, vomiting, anosmia and ageusia. Significant symptoms persisted and a second nasopharyngeal swab test for SARS CoV-2 RNA performed 37 days after the onset of symptoms was also positive. Most symptoms gradually resolved, and around the middle of May, she still complained of mild headache and asthenia.

Case 2 is another woman, also in her 50’s, who by the beginning of May began experiencing fever, headache, sore throat, cough, asthenia, rhinorrhea, arthralgia, myalgia and nausea. She contacted the São Caetano platform and a nasopharyngeal swab test for SARS CoV-2 RNA was positive (5 days after the onset of symptoms). Her symptoms persisted and a second nasopharyngeal swab test for SARS CoV-2 RNA performed 24 days after the onset of symptoms remained positive. She remained symptomatic, about 35 days after the onset of symptoms.

Since symptoms were relatively mild, both women were advised to remain at home. They did not undergo additional testing or received any treatment and there was a gradual improvement in their clinical condition.

Due to the persistence of symptoms and a prolonged positive RT-PCR result, it was decided to investigate the replicative capacity of their SARS-CoV-2 virus. Swab samples obtained at day 37 (case 1) and day 24 (case 2) were inoculated into Vero CCL81 cells and diagnostic tests were performed on the cell culture supernatant and intracellular fractions, as described below.

### Virus Identification: RNA extraction, PCR amplification and viral culture

All specimens were handled according to laboratory biosafety guidelines. Nasopharyngeal samples were subjected to total nucleic acid extraction with the QIAamp viral RNA kit (QIAGEN, Hilden, Germany), according to the manufacturer instructions. Samples were then subjected to RT-PCR (RealStar® SARS-CoV-2 RT-PCR Kit 1.0, *Altona Diagnostics)* followed by DNA amplification (Roche LightCycler® 96 System).

Viral culture for SARS-CoV-2, conducted in a biosafety level-3 facility, utilized Vero CCL81 cells (ATCC® CCL-81™) in Dulbecco minimal essential medium supplemented with 10% heat-inactivated fetal bovine serum and antibiotics/antimycotics .Nasopharyngeal samples were inoculated into a Vero cell culture in plastic bottles (Jet biofilm, 12,5 cm^2^ area, 25 mL capacity) and incubated in a 37°C incubator in an atmosphere of 5% CO_2_. Cultures were maintained for at least 2 weeks and observed daily for evidence of cytopathic effects (CPEs). At least two subcultures were performed on each sample. The detection of CPEs was investigated using an inverted microscope (Nikkon, Japan) and the presence of virus in supernatants from cultures showing CPEs was determined by specific RT-PCR, as described above. RT-PCR analysis was performed using RNA extracted from culture supernatants obtained two passages after the initial inoculation.

### Ultrastructural examination

A standard quantity of cells from the culture flasks inoculated with samples from patients 1 and 2, after at least two passages, were transferred to a 1.5 ml centrifuge tube containing 1.2 ml 3% glutaraldehyde in phosphate-buffered saline (PBS) at pH 7.4. A subsequent fixation occurred in a mixture of 1 vol 3% osmium tetroxide in PBS plus 1 vol aqueous 3% potassium ferrocyanide. Dehydration was performed by emersion in a series of increasing ethanol concentrations and 100% acetone. Embedding was in LX Epon. Ultrathin sections were obtained with an Ultracut microtome. Observations were carried out in a 20-20 Jeol Electron microscope.

## Results

Cytopathic effects were observed in the Vero cell cultures incubated with samples from both patients after three passages (Fig. 1B) and the presence of replicating SARS-CoV-2 in culture supernatants was confirmed by real-time RT-PCR.

**Figure 1.**
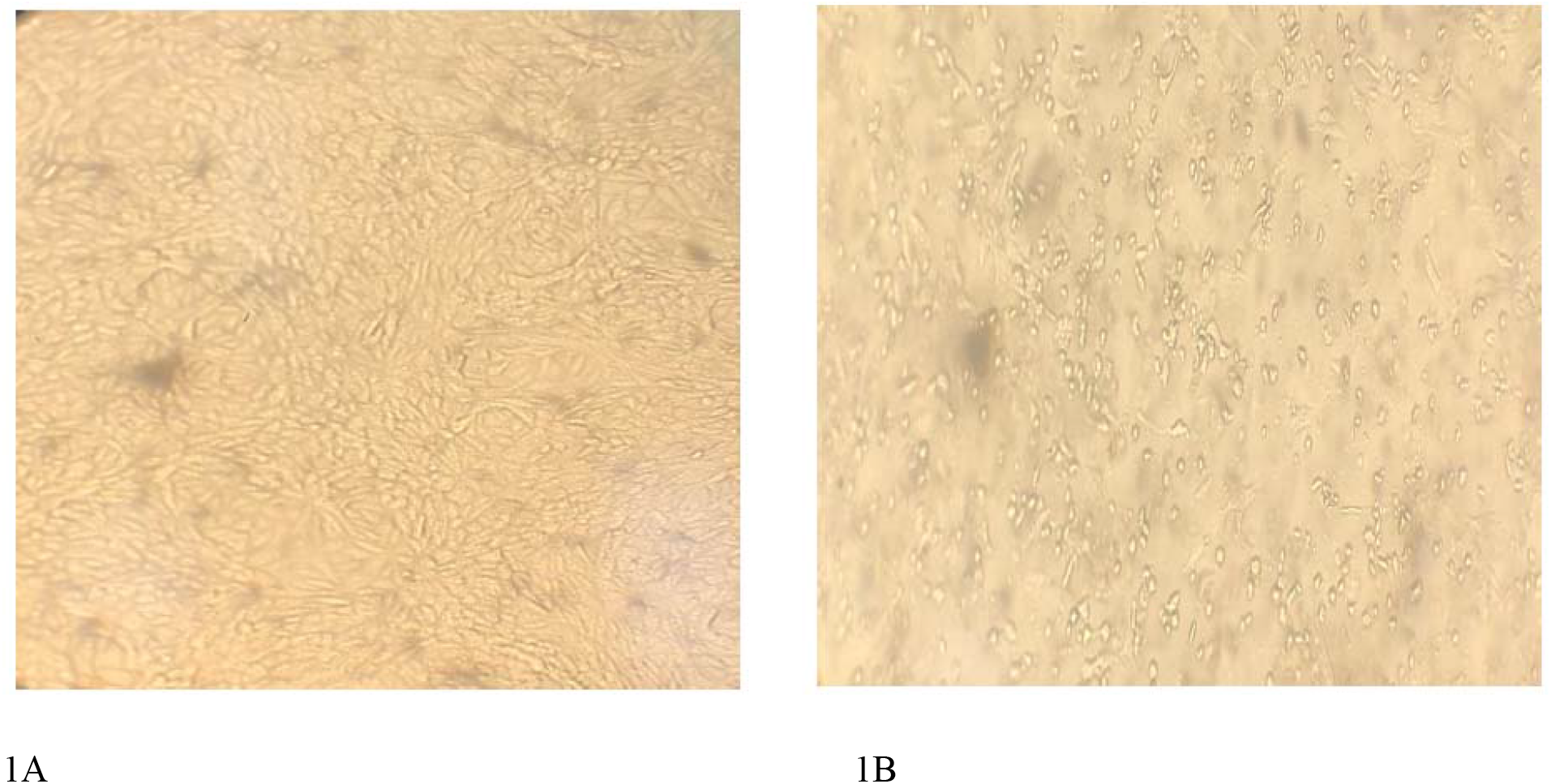
Induction of cytopathic effects on Vero cells after inoculations with nasopharyngeal swabs from women with SARS-CoV-2. Cultured Vero cells were untreated (A) or inoculated with material from nasopharyngeal swabs of patients positive for SARS-CoV-2 (B).

In addition, by electron microscopy, aggregates of elongated and spheroid particles ranging in size from around 60 nm to140 nm with peripheral spike-like projections consistent with the morphology described for SARS-CoV-2 (9) were observed (Fig. 2). The major and minor axes of the virus profiles were 100 and 58 nm, respectively. Measurements of the orthogonal long and short axes of several virus particles, located close to two cells in the same preparation had the following mean dimensions with the corresponding standard error of the mean, respectively, 90 ± 4.5 nm (n=22) and 62 ± 5.1 nm (n=22). Viral particles were seen mainly at the cell periphery and eventually inside cytoplasmic vacuoles (Fig. 2).

**Figure 2.**
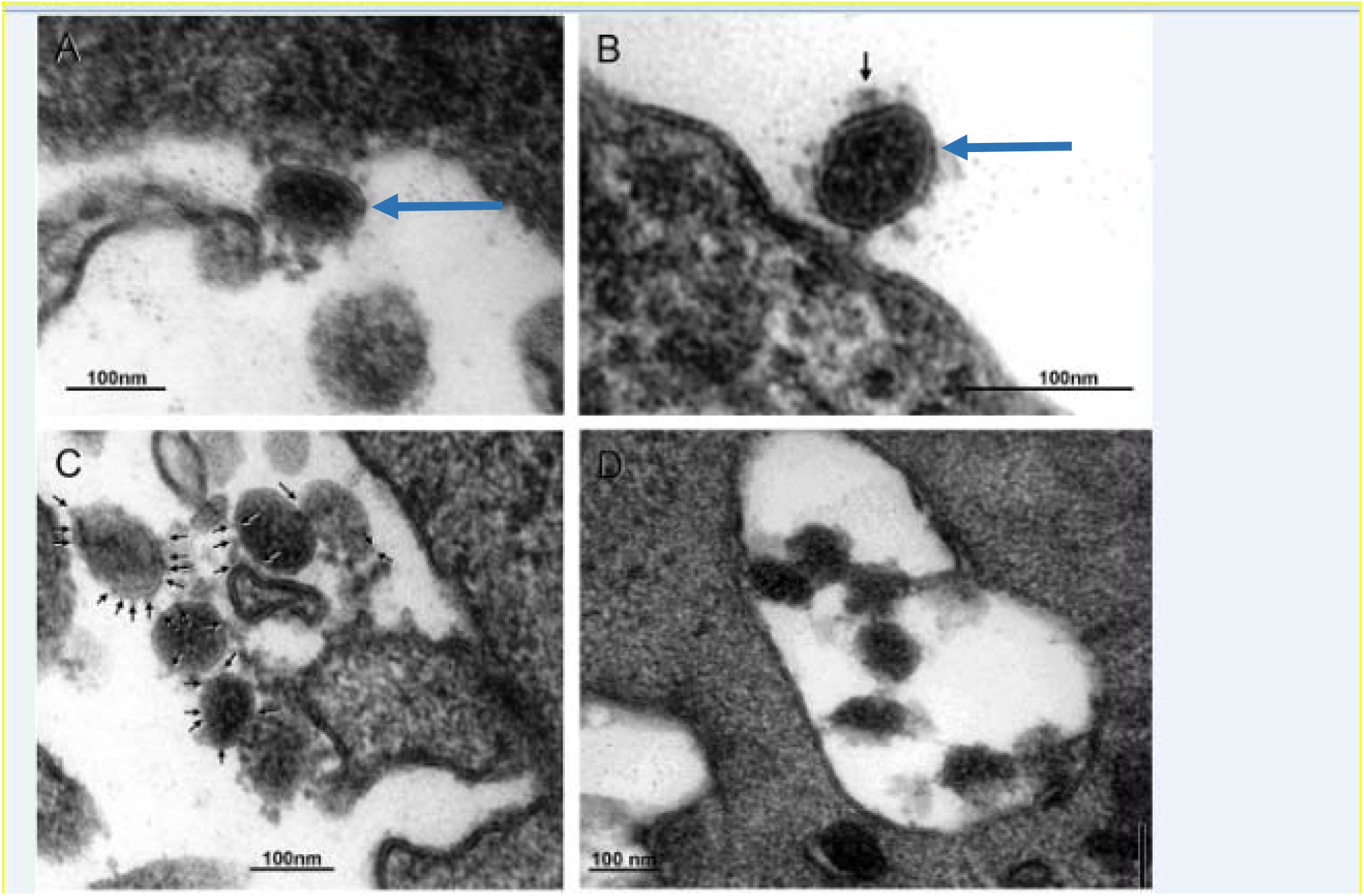
Electron microscopy of Vero cells inoculated with nasopharyngeal samples from women infected with SARS-CoV-2. Panel A, B, C, D are representative thin section electronmicrographs of the detection of SARS-CoV-2. Legend to panels A, B, C, D. Long blue arrows indicate elongated and spheroid viral particles, respectively, attached to the cell border membrane in panels A and B. In panel B small arrow points to a virus spike. In panel C small arrows indicate petite and longer virus spikes. Panel D contains several viral particles inside a cytoplasmic vacuole.

## Discussion

Two women positive for SARS-CoV-2 presented with flu-like symptoms that persisted for a longer time than is typical. This led to the collection of a second nasopharyngeal swab at 24 and 37 days, respectively, after symptom onset that resulted in the identification of replication-competent virus in both women. To the best of our knowledge, there are no previous reports of replication-competent virus being isolated three weeks after symptom onset. The cases described here also indicate that replication-competent virus may persist in mildly symptomatic patients who do not require hospitalization for a prolonged time period.

According to WHO updated recommendations on the criteria for discharging SARS-CoV-2-positive individuals from isolation, patients must be clinically recovered (symptom-free) (10). Our data reinforce that even mildly symptomatic individuals are potentially contagious. Recently, there have been descriptions of individuals who initially tested positive for SARS-CoV-2 RNA, became virus-negative but subsequently again became PCR-positive (11,12). This may be due to either reinfection following exposure to another infected person or by reactivation of latent virus (13). Reinfection, latent virus reactivation and prolonged viral shedding may represent unique presentations of this infection in different patients or, alternate phases of the same infection. Immunological and clinical characteristics of individual patients, as well as genomic characteristics of the involved viral strains may help determine the natural history of COVID-19 and the different phases of disease, as described above.

Further clarification of the frequency of presumed prolonged infectivity, as illustrated by the cases described in this communication, will be defined by prospective follow-up studies involving a greater number of individuals. Nevertheless, this report highlights that individuals with prolonged but mild symptoms can remain positive for replication-competent virus, highlighting the need for such individuals to exercise appropriate precautions to avoid potential transmission of SARSCoV-2 in their community.

## Data Availability

All data referred to in the manuscript is avaiable for consultation at any time.

## Declaration of interests

The authors declare that they have no known competing financial interests.

## Notes

### Competing Interest Statement

The authors have declared no competing interest.

### Funding Statement

Authors involved in the manuscript or study design did not receive any kind oy payment for their service.
The study was funded by Virology Laboratory from Instituto de Medicina Tropical-Faculdade de Medicina da Universidade de Sao Paulo.

### Author Declarations

The study was approved by the local ethics committee (Comissao de Etica para Analise de Projeto de Pesquisa - CAPPesq, protocol No. 13915, dated June 03, 2020).

## References

1 He X, Lau EHY, Wu P, Deng X, Wang J, Hao X, Lau YC, Wong JY, Guan Y, Tan X, Mo X, Chen Y, Liao B, Chen W, Hu F, Zhang Q, Zhong M, Wu Y, Zhao L, Zhang F, Cowling BJ, Li F, Leung GM. Temporal dynamics in viral shedding and transmissibility of COVID-19. Nat Med. 2020 May;26(5):672–675. doi: 10.1038/s41591-020-0869-5.

2 COVID-19 Investigation Team. Clinical and virologic characteristics of the first 12 patients with coronavirus disease 2019 (COVID-19) in the United States. Nat Med. 2020 Jun;26(6):861–868. doi: 10.1038/s41591-020-0877-5. Epub 2020 Apr 23. PMID: 32327757.

3 Wölfel R, Corman VM, Guggemos W, et al. Virological assessment of hospitalized patients with COVID-2019. Nature. 2020;581(7809):465–469. doi:10.1038/s41586-020-2196-x

4 Jeong HW, Kim SM, Kim HS, et al. Viable SARS-CoV-2 in various specimens from COVID-19 patients. Clin Microbiol Infect. 2020;S1198-743X(20)30427-4. doi:10.1016/j.cmi.2020.07.020

5 Liu WD, Chang SY, Wang JT, et al. Prolonged virus shedding even after seroconversion in a patient with COVID-19. J Infect. 2020;81(2):318–356. doi:10.1016/j.jinf.2020.03.063

6 van Kampen JJA. Shedding of infectious virus in hospitalized patients with coronavirus disease-2019 (COVID-19): duration and key determinants

7 Bullard J, Dust K, Funk D, et al. Predicting infectious SARS-CoV-2 from diagnostic samples Clin Infect Dis. 2020;ciaa638. doi:10.1093/cid/ciaa638

8 Arons MM, Hatfield KM, Reddy SC, et al. Presymptomatic SARS-CoV-2 Infections and Transmission in a Skilled Nursing Facility. N Engl J Med. 2020;382(22):2081–2090. doi:10.1056/NEJMoa2008457

9 Feng W, Zong W, Wang F, Ju S. Severe acute respiratory syndrome coronavirus 2 (SARS-CoV-2): a review. Mol Cancer. 2020 Jun 2;19(1):100. doi: 10.1186/s12943-020-01218-1. PMID: 32487159; PMCID: PMC7264920.

10 World Health Organization. Clinical management of COVID-19 (Interim Guidance) https://www.who.int/publications-detail/clinical-management-of-covid-19, published 27 May 2020.

11 Liu F, Cai ZB, Huang JS, Yu WY, Niu HY, Zhang Y, Sui DM, Wang F, Xue LZ, Xu AF. Positive SARS-CoV-2 RNA recurs repeatedly in a case recovered from COVID-19: dynamic results from 108 days of follow-up. Pathog Dis. 2020 Jun 1;78(4):ftaa031. doi: 10.1093/femspd/ftaa031. PMID: 32592396; PMCID: PMC7337794.

12 Li Y, Ji D, Cai W, Hu Y, Bai Y, Wu J, Xu J. Clinical characteristics, cause analysis and infectivity of COVID-19 nucleic acid repositive patients: A literature review. J Med Virol. 2020 Sep 5. doi: 10.1002/jmv.26491. Epub ahead of print. PMID: 32890414.

13 Goldman JD, Wang K, Roltgen K, Nielsen SCA, Roach JC, Naccache SN, Yang F, Wirz OF, Yost KE, Lee JY, Chun K, Wrin T, Petropoulos CJ, Lee I, Fallen S, Manner PM, Wallick JA, Algren HA, Murray KM, Su Y, Hadlock J, Jeharajah J, Berrington WR, Pappas GP, Nyatsatsang ST, Greninger AL, Satpathy AT, Pauk JS, Boyd SD, Heath JR. Reinfection with SARS-CoV-2 and Failure of Humoral Immunity: a case report. medRxiv [Preprint]. 2020 Sep 25:2020.09.22.20192443. doi: 10.1101/2020.09.22.20192443. PMID: 32995830; PMCID: PMC7523175.

